# Leveraging the wearable 1-lead ECG signal: From cardiac rhythm to cardiac function assessment

**DOI:** 10.64898/2026.02.02.26345091

**Authors:** Viktor van der Valk, Douwe Atsma, Roderick Scherptong, Marius Staring

## Abstract

The electrocardiogram (ECG) is a critical tool in the diagnosis and monitoring of cardiovascular disease. Although traditional 12-lead ECGs offer comprehensive insights into the electrical activity of the heart, they typically require clinical settings and expert interpretation, which limits their accessibility. In contrast, smartwatch 1-lead ECGs can be recorded at home, allowing more frequent and rapid monitoring. This opens opportunities not only for early detection but also for enhancing patient autonomy. This study investigates whether 1-lead ECGs can provide information beyond heart rhythm, specifically whether they can be used to assess left ventricular function (LVF) using explainable deep learning models. Our findings show that LVF can be accurately predicted from 1-lead ECGs (AUC = 0.883), nearly matching the performance of 12-lead ECGs (AUC = 0.897). These results suggest that 1-lead ECGs, when combined with interpretable AI, could support broader clinical applications and empower patients, particularly in resource-limited or remote settings.

## I Introduction

Cardiovascular disease remains the leading global cause of death, underscoring the need for accessible and accurate diagnostic tools. The 12-lead electrocardiogram (ECG) is the clinical gold standard for assessing cardiac electrical activity and diagnosing a wide range of heart conditions. However, its application is largely confined to clinical settings due to the need for specialized equipment and trained personnel.

In contrast, the 1-lead ECG offers a portable and cost-effective alternative. Devices such as smartwatches and chest patches enable patients to record ECGs independently, facilitating continuous and remote monitoring. However, the reduced spatial resolution of 1-lead ECGs limits their diagnostic scope, particularly for detecting structural abnormalities. These devices have primarily been optimized for rhythm analysis, especially atrial fibrillation (AF) detection, with several FDA- and EMA-approved systems (e.g., Apple Watch, KardiaMobile) achieving high accuracy in real-world applications^1,2,3^.

Deep learning has significantly advanced the interpretability and accuracy of ECG-based diagnostics^4,5^. In the 2017 PhysioNet Challenge, top-performing AF classifiers used convolutional and recurrent neural networks for 1-lead ECG analysis^6,7^. Since then, explainable AI methods have enabled more transparent and trustworthy models for AF and other arrhythmias^8,9,10^, supporting broader adoption in clinical practice.

Beyond arrhythmias, recent work has extended 1-lead ECG applications to detecting heart failure^11^, chronic kidney disease^12^, and age-related cardiac changes^13^. Moreover, predictive modeling using individual leads, such as Lead I or II, has shown potential for detecting myocardial infarction, left ventricular dysfunction, and myocardial injury, although typically with reduced accuracy compared to full 12-lead recordings^14,15,16,17^. These efforts illustrate the growing interest in the use of reduced-lead ECGs for not only rhythm assessments, but also structural and functional cardiac assessments.

Despite these advances, few studies have directly compared wrist-based 1-lead ECGs with full 12-lead ECGs acquired from the same patients. Most prior work has either focused on specific conditions (e.g., arrythmias) or used leads derived from 12-lead datasets, limiting their applicability to real-world wearable recordings. Moreover, while arrhythmia detection has dominated the use of 1-lead ECGs, their potential to identify structural cardiac changes, such as ventricular dysfunction, remains underexplored, although such abnormalities often leave subtle but detectable signatures in the electrical signal. Attia *et al*. (2022) demonstrated that left ventricular dysfunction (LVD) could be predicted from a 1-lead smartwatch ECG with reasonable accuracy (AUC = 0.885) in a general cardiac patient population^18^. However, to our knowledge, left ventricular function (LVF) assessment from 1-lead smart-watch ECGs has not yet been evaluated with explainable AI, nor in the post–myocardial infarction setting, a population in which early detection of ventricular dysfunction is particularly relevant. The post-myocardial infarction population is homogeneous, since scar tissue is the main cause of abnormalities. The confined heterogeneity in this population is due to the location and severity of myocardial infarction. This is an important aspect that can be leveraged in explainable AI models.

This study conducts a head-to-head comparison of wrist-based 1-lead ECGs, recorded at home as part of the follow-up protocol, and standard 12-lead ECGs, matched by patient and acquisition time, in individuals recovering from myocardial infarction. Focusing on cardiac morphology, we apply deep learning and explainable AI to evaluate whether wearable 1-lead ECGs can move beyond rhythm analysis and provide clinically meaningful insight into structural cardiac abnormalities.

## II Methods

### II.A Data

The study uses two datasets, a 1-lead ECG dataset and a 12-lead ECG dataset from the same cohort, see Figure 1. All data was obtained from patients who were diagnosed with acute coronary syndrome, between 2018 and 2023 and treated at the Leiden University Medical Center in the Netherlands. The 1-lead dataset contains 63.950 raw 30-second, 1-lead ECG signals recorded at a frequency of 300Hz. These signals were recorded by 708 patients (77.3% male, age 59.5 ± 10.8 years). The 1-lead ECG signals were recorded at home with a Withings ECG Smartwatch as part of a clinical telemonitoring program^19,20^. The 12-lead dataset contains 77.005 raw 10-second, 12-lead ECG signals recorded at a frequency of 500Hz. These signals are obtained from 6.703 patients (76.3% male, age 60.5 ± 11.3 years). The study protocol, identified as nWMODIV2 2022006, received approval from the institutional review board (niet-Wet medische wetenschappelijk onderzoek (nWMO) commissie Divisie 2), which waived the requirement to obtain informed consent for the use of ECG data of individual pseudonymized patients.

**Figure 1.**
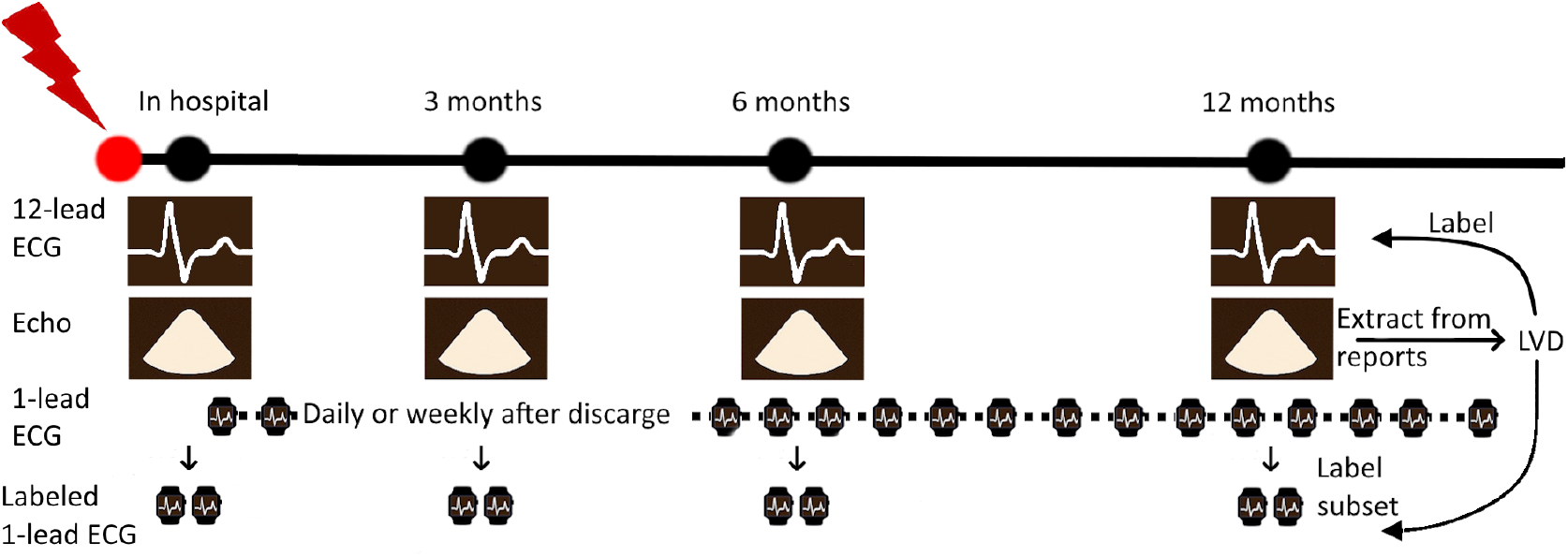
Clinical monitoring and data collection: Timeline indicates the 12-lead ECG, 1-lead ECG and echocardiogram data collection following the clinical protocol. 12-lead ECGs and echocardiograms are collected during hospital visit as part of the standard monitoring protocol after MI. 1-lead ECGs are collected at home by patients on suggested intervals after discharge. A subset of the 1-lead ECGs, when acquisition overlaps with echocardiography, is labeled with the LVF label extracted from the echocardiography reports.

### II.B Data preprocessing

Preprocessing of both 1-lead and 12-lead ECG signals followed the procedure described in Van der Valk *et al*. (2025)^9^, see Figure 2 for an overview. The raw ECG signals were segmented into individual heartbeats, each defined as an 800 ms interval centered on the R-peak, 400 ms preceding and 400 ms following. R-peak detection was performed using a method inspired by RPNet, a U-Net–structured CNN, previously trained on manually annotated peak locations as described in Vijayarangan *et al*. (2020)^21^.

**Figure 2.**
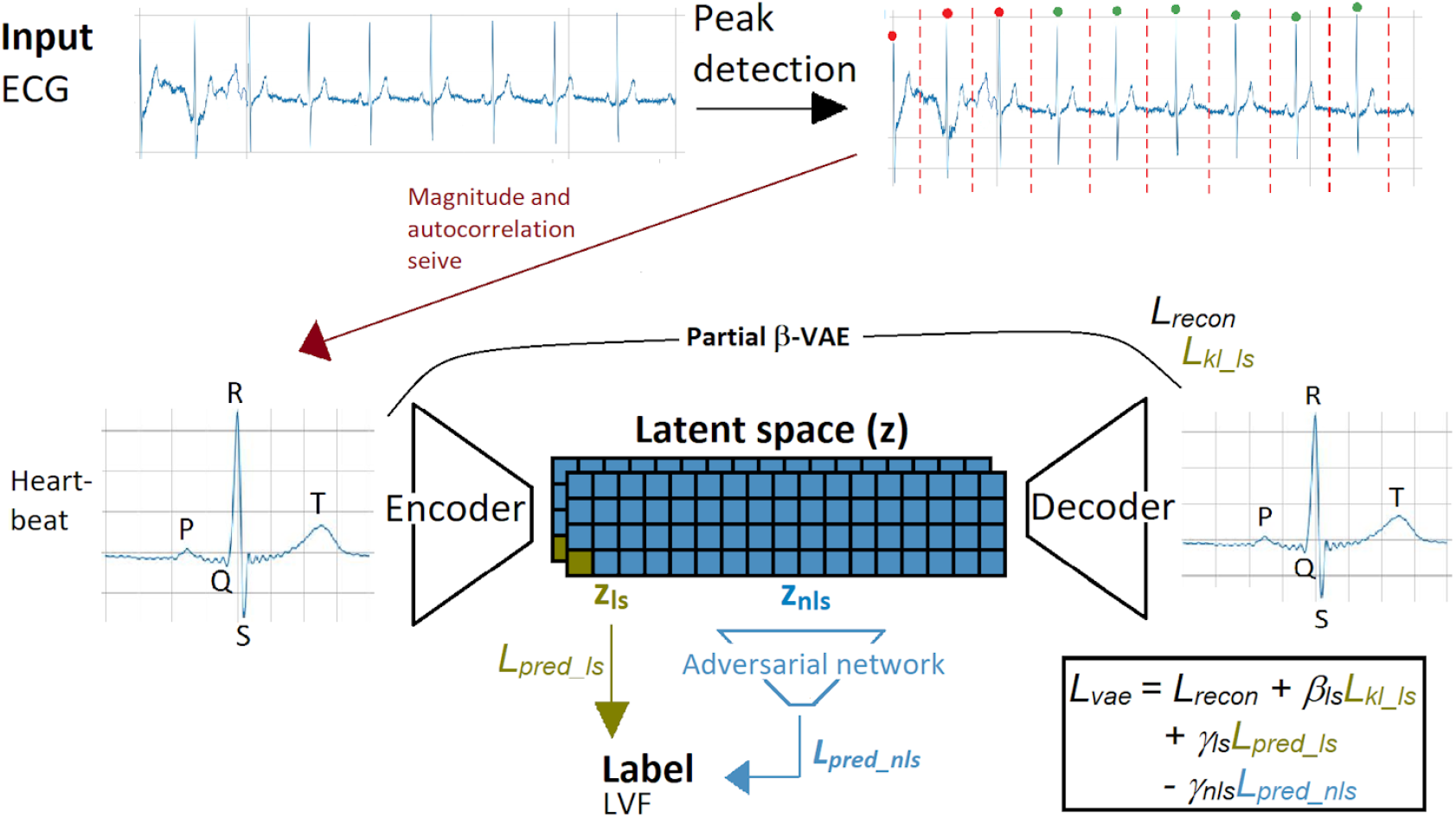
Overview of our Processing and Prediction Framework: Initially, the 12-lead ECGs are split into individual heartbeats using RPNet^21^. Subsequently, the heartbeats are sieved in two stages: the first based on individual magnitude, identifying low magnitude as indicative of a flat signal, and the second utilizing autocorrelation to assess correlation with other heartbeats from the same signal. The remaining heartbeats (indicated with green dots) are used as input for the VAE. The encoded representation *z* is derived, comprising *z*_nls_ and *z*_ls_. While *z*_ls_ directly contributes to LVF label prediction with loss *L*_pred_ls_, *z*_nls_ serves as input for an adversarial MLP also predicting the LVF label, introducing loss *L*_pred nls_. The decoder’s input is sampled from *z* to produce the reconstructed heartbeat, subsequently compared with the original input heartbeat, resulting in the reconstruction loss *L*_recon_.

Because the 1-lead recordings were 30s in duration compared to the 10s duration of the 12-lead recordings, the 1-lead data yielded a greater number of segmented beats. Given the low intra-recording variability and to maximize the available training data for the 1-lead model, all valid heartbeats were retained for this modality. Segmented beats containing excessive noise or flatline segments were excluded; for the 12-lead case, exclusion was done if any lead exhibited such artifacts.

### II.C Left ventricular function

We selected LVF as a prediction target due to its reflection of structural changes after MI and the presence of well-characterized ECG features. Several of these, such as the first QRS integral, ST integral, ST–T integral, and ST–T area, have shown discrimination in recovering MI patients^22^. The predictive capacity of these features differed per lead and MI location^23^. T-wave morphology changes, including inversion and subsequent reinversion, have also been linked with favorable LVF recovery^24^.

The ECG signals were labeled with LVF impairment, consisting of 4 ordinal categories: normal function, mild impairment, moderate impairment, and severe impairment. Labeling was performed using large language model (LLM)–based classification of echocardiogram reports obtained as part of the routine clinical workflow^25^. An echocardiogram report was used as a label if the examination was conducted within seven days before or after the corresponding ECG recording. The reports themselves were based on visual interpretation of the echocardiograms by clinicians.

### II.D Model overview and training

LVF prediction from both 1-lead and 12-lead ECGs employed the same model and training strategy as succesfully employed in Van der Valk *et al*. (2025). A partial *β*-variational autoencoder (VAE) (Figure 2) was used for both modalities. The architecture consisted of seven recurrent convolutional blocks in both the encoder and decoder. Four separate model instances were trained: i. a 12-lead model, a model trained and evaluated on the 12-lead ECGs, ii. a lead I model, a model trained and evaluated on lead I of the 12-lead ECGs, iii. a pretrained 1-lead model, a model pretrained on lead I of the 12-lead ECG and finetuned and evaluated on the 1-lead ECGs, and iv. a 1-lead model, a model trained and evaluated on the 1-lead ECGs. The models expect an input of shape 12 × 400; therefore, for the 1-lead recordings, each 1 × 400 heartbeat was duplicated along the lead dimension to match the required input shape.

### II.E. Feature visualization

The model output and interpretability is shown by means of attribute manipulation as explained in Van der Valk *et al*. (2025)^9^, see Figure 2. Each ECG heartbeat was encoded into *z*_nls_ and *z*_ls_, of dimension 599 and one respectively. Then *z* was decoded to reconstruct the signal. For attribute manipulation 6 hypothetical signals were created by replacing *z*_ls_ with six values: four means for the four LVF groups (*S*_1_–*S*_4_) and two extremes (*S*_0_: mean minus one standard deviation of severely impaired group; *S*_5_: mean plus one standard deviation of normal LVF group). These six patient-specific hypothetical signals were used to illustrate how ECG morphology changes across LVF impairment levels and which features the model leverages for classification. These signals create a context for the ECG heartbeat w.r.t. LVF, allowing comparison against the whole database. This facilitates signal interpretation and potentially enable interpretation by less trained medical personnel.

## III. Experiments and Results

### III.A. Experiments

Both datasets were split into a training set (89% of signals) and a holdout test set (11% of signals). The test set was constructed exclusively from the overlapping portion of the datasets, where both patients and acquisition dates (within a seven-day window) were overlapping, to enable a head-to-head comparison. A 5-fold cross-validation was performed in both training sets, maintaining an 85:15 ratio between the training and validation set within each fold. All data splits were grouped by patient and stratified by label when applicable. Training continued until convergence, defined as no improvement in validation loss for 15 consecutive epochs. To mitigate overfitting, we employed balanced sampling, early stopping, L2 regularization on the fully connected layer, and weight decay with the Adam optimizer. To prevent gradient explosion, He initialization was used^26^, and to avoid numerical overflow, the standard deviations of the posterior *q*(*z*|*X*) were clipped to the interval [-10,3] prior to sampling. The models were implemented and trained on a 24Gb GPU. Hyperparameter tuning resulted in *β*_*ls*_ = 0, *β*_*nls*_ = 0.001 and *γ* = 2.0 for both models. This is in line with the experiments shown by Van der Valk *et al*. (2025)^9^.

### III.B. Feature evaluation

LVF predictions were evaluated using three classification metrics: the Area Under the Receiver Operating Characteristic Curve (AUROC), average precision score (AP) and a heart-beat consistency score (HBC). These metrics were applied to a binary classification task (positive class: severe/moderate impairment vs. negative class: mild impairment/normal function), reflecting clinically relevant groupings and enabling comparison with previous studies. The HBC score was defined as the portion of heartbeats from the same signal that was classified in the same group. The AUROC and AP scores were calculated with the averaged predictions from all heartbeats in a signal.

Heartbeat reconstruction performance was assessed using the mean squared error (MSE) between the ECG input and the output signal. For intermodel comparisons of prediction scores and MSE, paired t-tests were performed. The confidence intervals (95%) for AUC, AP and HBC scores were estimated by bootstrapping with n = 1000 samples of the combined output distribution (5 models). The confidence interval for MSE was estimated directly with the combined output error distribution.

## III.C. Results

7.118 1-lead ECG signals and 7.593 12-lead ECG signals were be labeled with LVF. Respectively, 14% and 11% of these were labeled with severe/moderate impaired LVF.

Several data quality issues were identified in the 1-lead recordings, including devices worn on the wrong wrist, leading to signal inversion, device sharing among multiple individuals, and acquisition during non-resting states. These factors introduce substantial noise and may impair model performance. To mitigate this, we applied an autocorrelation-based filter that utilized preceding and subsequent signals from the same patient to exclude low-quality recordings. After filtering, 5,480 1-lead signals comprising 108,364 heartbeats from 460 patients were retained for model training. For the 12-lead recordings, no comparable quality concerns were observed; 7,197 signals and 19,933 heartbeats from 1,856 patients were included. The corresponding test sets consisted of 900 1-lead signals and 559 12-lead signals from 144 patients.

Table 1 and Figure 3a and 3b summarize the performance of the four models for LVF prediction on the test set. As anticipated, the full 12-lead model achieved the highest predictive performance across all metrics. The 1-lead models demonstrated reasonable discriminative ability, with only a modest reduction in performance compared to the 12-lead model. Pretraining the 1-lead model using lead I of the 12-lead ECG improved AUC and AP but did not improve heartbeat consistency (HBC). The lead I model exhibited the lowest AUC yet ranked second HBC after the full 12-lead model. In Figure 3a and 3b sensitivity at 0.9 and 0.95 are indicated with grey dotted lines. For a sensitivity of 0.9 (modere/severe impaired LVF detected in 90% of cases), specificities are: 0.64, 0.43, 0.71, and 0.65 for respectively the 12-lead, lead I, pretrained 1-lead and 1-lead models; for a sensitivity of 0.95 the respective specificities are: 0.49, 0.50, 0.55, and 0.33. All models showed high intra-recording stability, as reflected by elevated HBC scores: most QRS complexes within a single recording were consistently assigned to the same binary class. This robustness underscores the reliability of signal-level predictions. Figure 4 illustrates the AUC when heartbeat-level predictions were aggregated by averaging within each signal.

**Table 1.** Comparison of all models for both reconstruction (MSE) and prediction of LVF (AUROC, AP and HBC). The AUROC, AP, and HBC scores are shown for the prediction of the moderate/severe impaired (positive) vs. normal/mild impaired groups. The AUROC, AP and HBC metrics show the bootstrapped mean (and 95% confidence interval) on the test set.

| Model | MSE ↓ | AUROC ↑ | AP ↑ | HBC ↑ |
| --- | --- | --- | --- | --- |
| 12-lead | 0.027<br>[0.010-0.087] | **0.897**<br>[0.887-0.906] | **0.600**<br>[0.573-0.626] | **0.991**<br>[0.857-1.000] |
| 12-lead<br>(Lead I) | 0.007<br>[0.002-0.036] | 0.805<br>[0.792-0.816] | 0.357<br>[0.332-0.382] | 0.980<br>[0.667-1.000] |
| 1-lead<br>(pretrained) | **0.004**<br>[0.001-0.011] | 0.883<br>[0.881-0.886] | 0.511<br>[0.501-521] | 0.959<br>[0.632-1.000] |
| 1-lead | **0.004**<br>[0.002-0.012] | 0.863<br>[0.859-0.866] | 0.459<br>[0.449-0.469] | 0.961<br>[0.647-1.000] |

**Figure 3.**
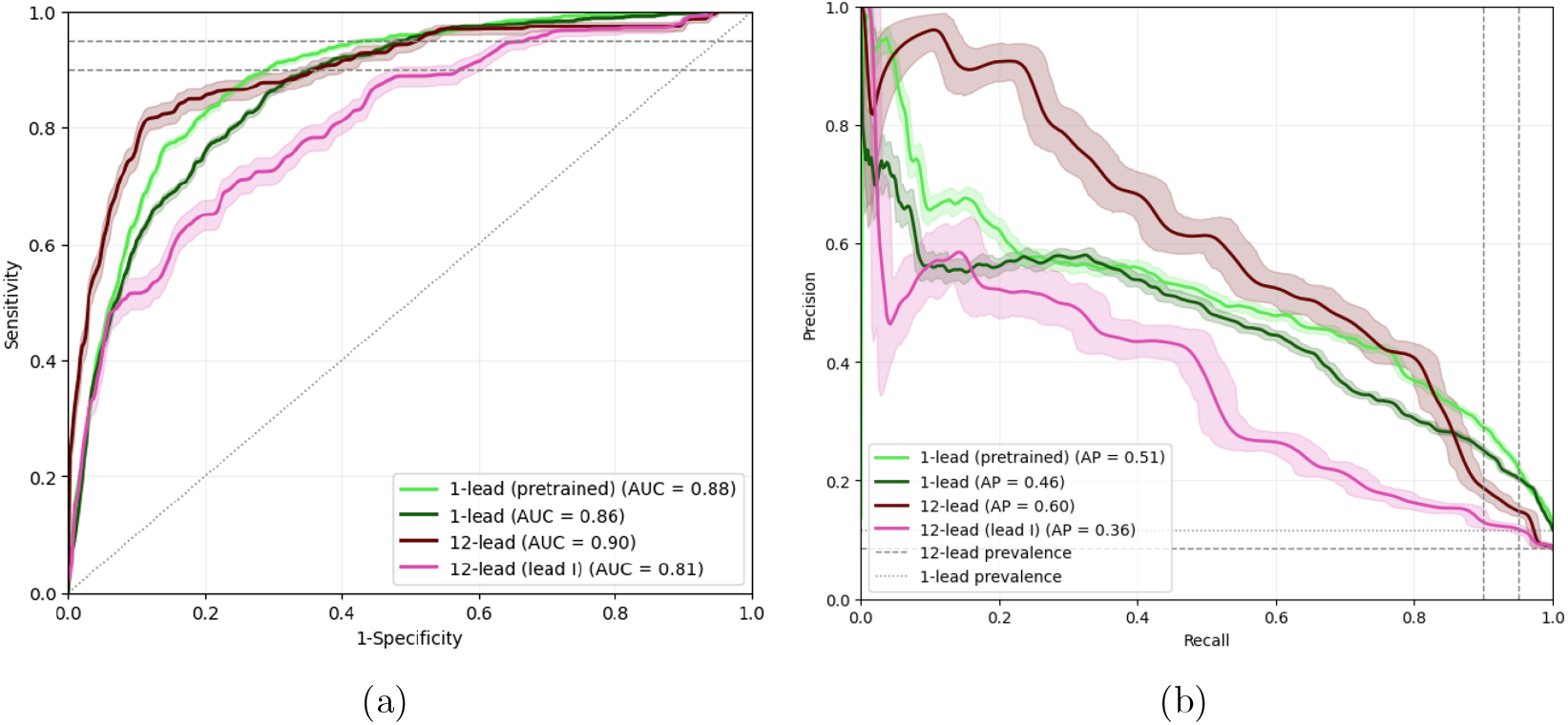
ROC and PR analysis including 95% confidence intervals, comparing the four models. (a) The ROC curves with the gray vertical dashed lines indicating recall/sensitivity 0.9 and 0.95. (b) The PR curves with the gray vertical dashed lines indicating recall/sensitivity 0.9 and 0.95. AP = average precision

**Figure 4.**
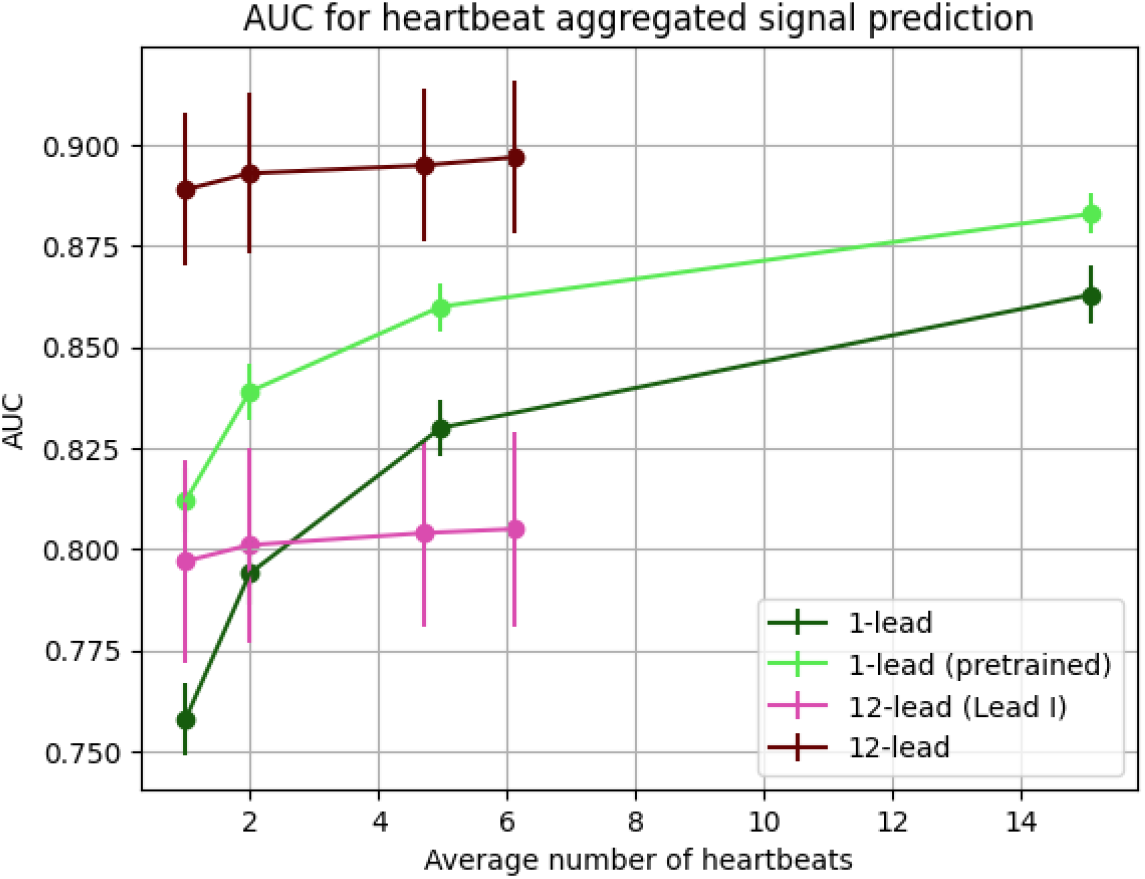
AUROC values for intra-signal aggregation of predictions. 1-lead models benefit significantly more from post-processing than 12-lead models.

### III.D. Evaluation of interpretability

Figures 5a and 5b show histograms of predicted LVF values for the four LVF groups, as obtained from the pretrained 1-lead and full 12-lead ECG models. Both models demonstrated the ability to distinguish between LVF classes, even though distributions show overlap, the means of the distributions show separation.

**Figure 5.**
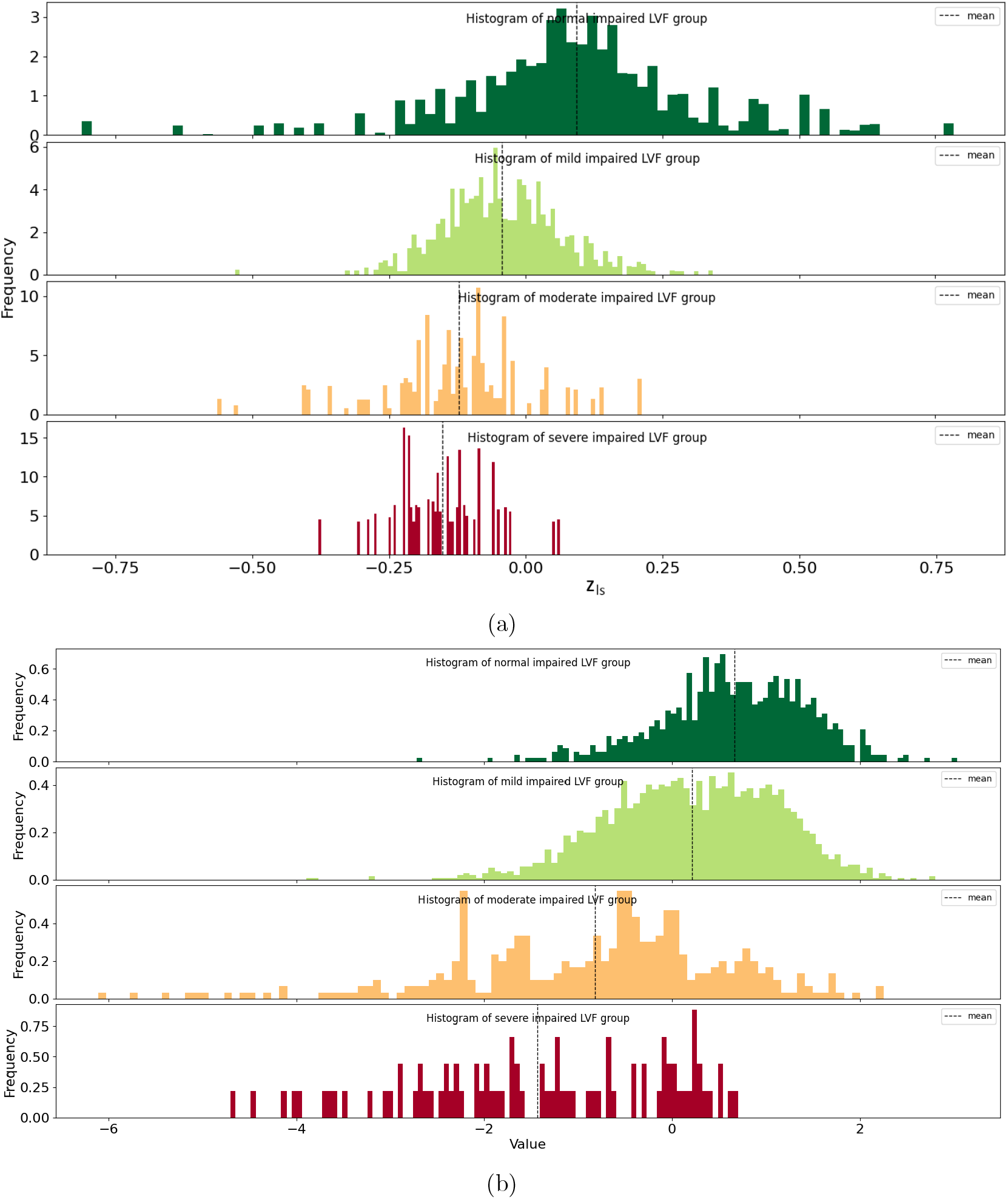
Histogram of the distributions of the continuous (a) 1-lead vs (b) 12-lead ECG-derived LVF values for the 4 LVF groups. The distributions show overlap, which indicates that the prediction is reasonable. The means of the distributions are indicated with a dotted vertical line.

Figure 6 and 7 show attribute manipulation of a heartbeat of the same patient with the pre-trained 1-lead and the full 12-lead model, respectively. The attribute manipulation can be interpreted as the rationale behind the model’s prediction. The 12-lead model shows a much more diverse set of features in multiple leads. Lead I of the 12-lead model shows relatively similar output to the 1-lead model.

**Figure 6.**
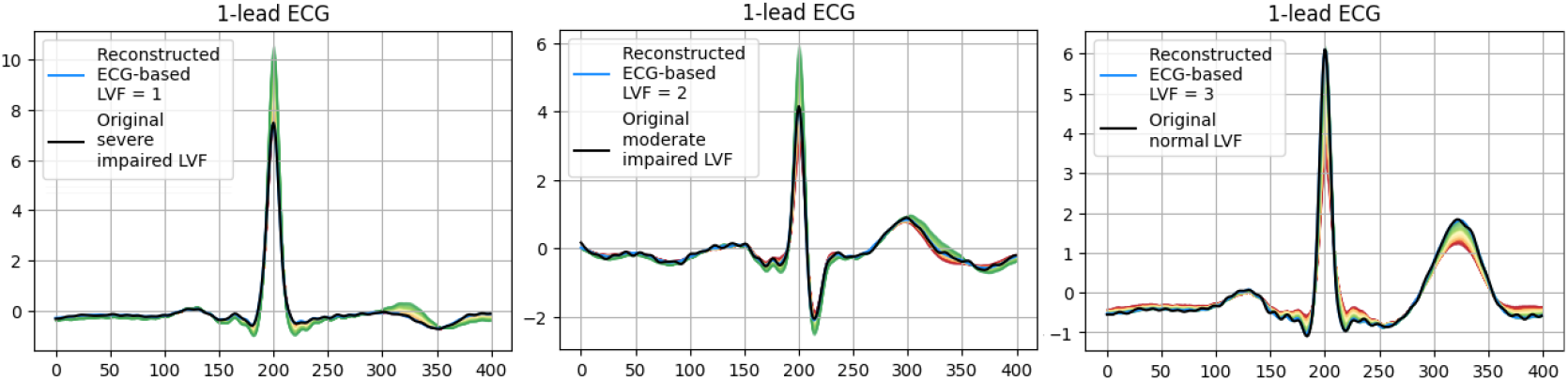
Attribute manipulation in three heartbeats with different LVF impairment. The black line shows the origin heartbeat of the 1-lead ECG, the blue line (largely covered by the black line here) shows the reconstructed heartbeat by the model. The green to red distributions around the reconstructed signal are the result of attribute manipulation and can be interpreted as the reason that the model classified the signal with the LVF value shown in the legend (0=severely impaired, 1=moderately impaired, 2=mildly impaired, 3=normal). The green signal shows the 1-lead signal that would be classified as having a normal LVF. The green signal in all figures shows a R-peak with larger amplitude, more pronounced Q- and S-waves and a T-wave with a slightly larger area. Both the T-wave and the R-peak are important prognostic factors in myocardial infarction. However, quantitative assessment of T-wave and R-peak changes after myocardial infarction is currently not part of the clinical routine in the non-acute setting.

**Figure 7.**
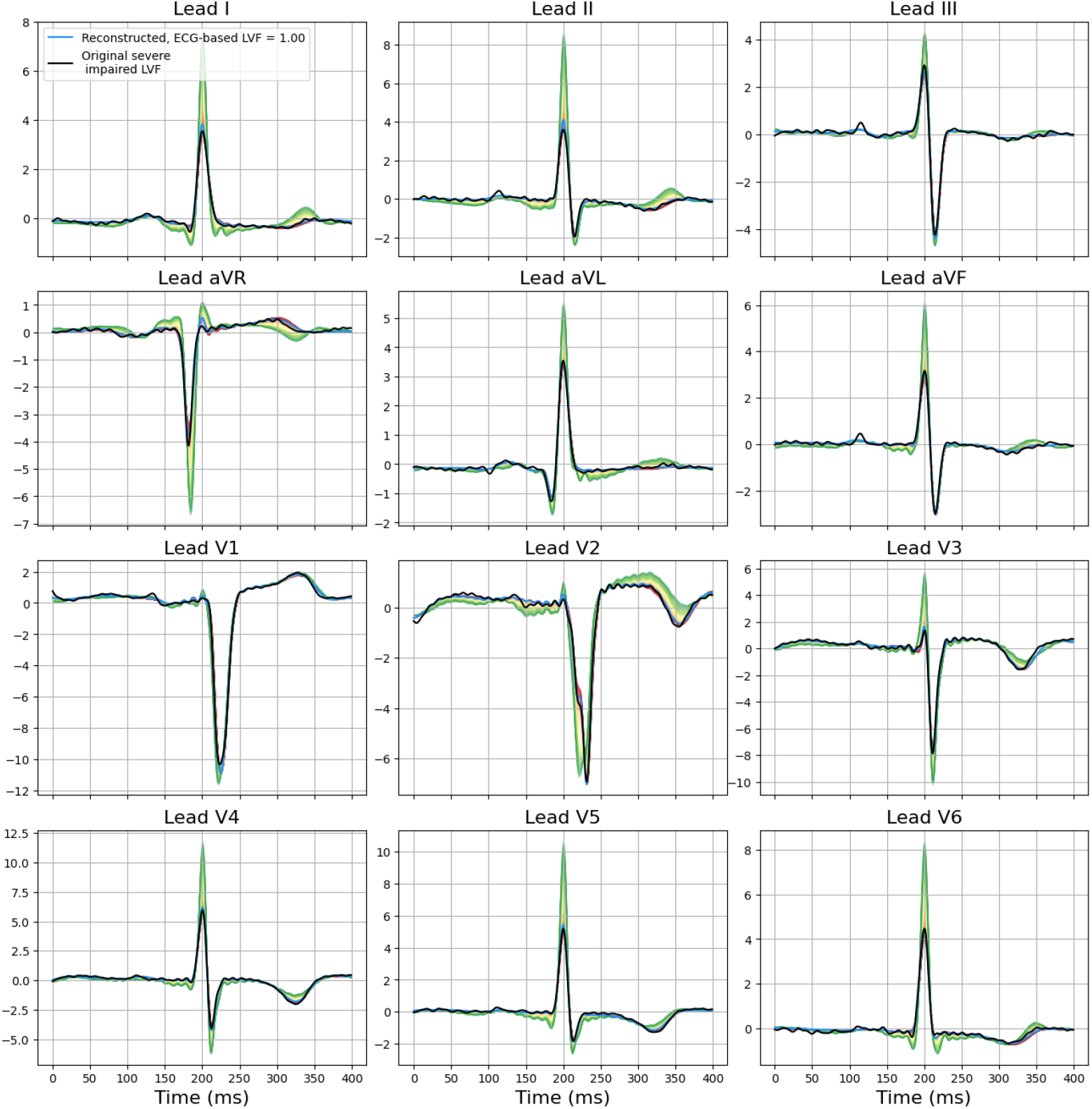
Attribute manipulation in a heartbeat with severe LVF impairment, the patient and visualization method are the same as in the first plot in Figure 6, see that caption for detailed explanation. In this case the green signal shows among others a larger R-peak amplitude, a T-wave with larger area and a R-peak without a notch in lead aVR, V2, aVF, V5, and V6. These are all important prognostic factors in myocardial infarction. However, quantitative assessment of these factors after myocardial infarction is currently not part of the clinical routine in the non-acute setting. Prediction to LVF: 0=severely impaired, 1=moderately impaired, 2=mildly impaired, 3=normal.

## IV. Discussion

This study provides a comparison of LVF prediction using matched 1-lead and 12-lead ECG datasets. As expected, the 12-lead model achieved the highest overall performance; however, the performance gap with the 1-lead models was modest, indicating that a substantial portion of LVF relevant information present in the full 12-lead ECG is also captured in a single lead. Model performance was comparable to, and in some instances slightly exceeded, previously reported results in broader cardiac populations^9,18^ (AUCs 0.83 and 0.89, respectively). Since early detection of ventricular dysfunction is particularly relevant in our post-myocardial infarction population, the findings of our study bear significant importance.

### Clinical utility and operating points

At a sensitivity of 0.90, specificity scores were 0.64 (full 12-lead), 0.43 (lead I), 0.71 (pretrained 1-lead), and 0.65 (1-lead). These operating points are usable in clinical pathways. The scores show the percentage of healthy patients that should not be reviewed for a test that correctly predicts left ventricular dysfunction with an accuracy of 0.9. They could be used, for example, to trigger manual review or flag patients as likely unchanged/low risk, especially in settings where 12-lead acquisition is not feasible. Notably, averaging heartbeat-level predictions within a recording preferentially improved the 1-lead models, consistent with (i) the larger number of heartbeats available in the longer 1-lead signal (30s vs. 10s) and (ii) noise cancellation through aggregation. Together with the observed high heartbeat consistency, this also supports the benefit of temporal aggregation over multiple recordings.

### Explainability as a driver for future research

A distinctive aspect of this work is the use of attribute manipulation to provide a transparent view of model reasoning. The 12-lead model leveraged a broader, multi-lead feature set (e.g. R-peak amplitude, notched R-peaks, T-wave morphology, see figure 7), whereas the 1-lead models used features that resembled the 12-lead feature set for lead I, which restricts interpretability and model performance, see figure 6. This observation raises important hypotheses for future investigation: (i) whether LVF prediction from a single lead is most effective in disease subtypes characterized by global or lateral abnormalities or lesions, where changes can be observed on lead I (and the 1-lead); (ii) whether multi-lead configurations are essential for more anterior or inferior infarctions; and (iii) how explainability can guide lead selection strategies for simplified ECG acquisition in wearable or resource-limited settings. The observed performance drop for the lead I model compared to the full 12-lead model reinforces the value of complementary spatial information, yet the improvement of the 1-lead model after heartbeat aggregation suggests that temporal averaging may partially compensate for reduced spatial coverage. Future work should validate these hypotheses and explore whether explainability-driven insights can inform adaptive acquisition protocols or subtype-specific prediction models.

Our model uniquely offers explainability that can facilitate clinical interpretation by less specialized personnel. By highlighting which waveform components drive predictions (e.g., T-wave morphology, R-peak amplitude, notched R-peaks), these tools can guide non-experts in verifying signal plausibility and understanding when predictions are likely reliable versus when additional expert review is warranted. This capability is critical for scaling LVF prediction into resource-limited settings and home-based care, where cardiology expertise may not be immediately available.

### Implications for wearables and remote care

While wrist-based 1-lead ECGs have primarily been optimized for rhythm analysis, such as atrial fibrillation detection, this study demonstrates their potential for structural and functional cardiac assessment. Although 1-lead ECGs are not intented to replace 12-lead ECGs, the combination of reasonable discriminative performance, transparent mechanism, and aggregation-driven stability, supports 1-lead ECGs as a practical tool for screening and potentially longitudinal monitoring. It could be used as a scalable clinical complement for (automated) early warning and for home-based follow-up where high quality 12-lead acquisition is not possible.

### Data quality and future work

We addressed 1-lead data quality challenges (device orientation errors, device sharing, non rest acquisition) using an autocorrelation based filter informed by adjacent recordings from the same patient. However, differentiating true pathological inversions from orientation-related inversions remains non-trivial, since several conditions transiently invert peaks or alter large ECG segments. We explored heartbeat autocorrelation, landmark based checks, and learned classifiers with mixed success. Future research directions to address these issues and extend currently methodology include: (i) acquisition protocol refinement to reduce incorrect device orientation and activity artifacts, (ii) robust automated quality control that combines signal-to-signal and even beat-to-beat consistency and anomaly detection, (iii) temporal modeling (e.g. multi visit aggregation) to convert stable beat-level consistency into patient-level trajectories, (iv) model performance analysis in clinical subtypes (global/lateral repolarization abnormalities vs. regional abnormalities) and (v) clinical validation with calibrated, use case specific thresholds (e.g. fixed 0.90 specificity), including health system impact metrics.

## Conclusion

The 12-lead ECG remains the gold standard for LVF prediction, yet 1-lead models achieve clinically meaningful performance, particularly when predictions are aggregated. Our model was notably designed to enable explainability of the predictions, highlighting a confined single-lead feature set in the 1-lead models versus multi-feature, multi-lead reasoning in the 12-lead model. This provides the mechanistic insight needed to scale these tools to real world applications.

## Data Availability

The data will not be made available due to privacy restrictions.

